# Workplace Contact Patterns in England during the COVID-19 Pandemic: Analysis of the Virus Watch prospective cohort study

**DOI:** 10.1101/2021.12.16.21267906

**Authors:** Sarah Beale, Susan Hoskins, Thomas Byrne, Wing Lam Erica Fong, Ellen Fragaszy, Cyril Geismar, Jana Kovar, Annalan M D Navaratnam, Vincent Nguyen, Parth Patel, Alexei Yavlinsky, Anne M Johnson, Robert W Aldridge, Andrew Hayward, on behalf of the Virus Watch Collaborative

## Abstract

**Background:** Workplaces are an important potential source of SARS-CoV-2 exposure; however, investigation into workplace contact patterns is lacking. This study aimed to investigate how workplace attendance and features of contact varied between occupations and over time during the COVID-19 pandemic in England.

**Methods:** Data were obtained from electronic contact diaries submitted between November 2020 and November 2021 by employed/self-employed prospective cohort study participants (*n*=4,616). We used mixed models to investigate the main effects and potential interactions between occupation and time for: workplace attendance, number of people in shared workspace, time spent sharing workspace, number of close contacts, and usage of face coverings.

**Findings:** Workplace attendance and contact patterns varied across occupations and time. The predicted probability of intense space sharing during the day was highest for healthcare (78% [95% CI: 75-81%]) and education workers (64% [59%-69%]), who also had the highest probabilities for larger numbers of close contacts (36% [32%-40%] and 38% [33%-43%] respectively). Education workers also demonstrated relatively low predicted probability (51% [44%-57%]) of wearing a face covering during close contact. Across all occupational groups, levels of workspace sharing and close contact were higher and usage of face coverings at work lower in later phases of the pandemic compared to earlier phases.

**Interpretation:** Major variations in patterns of workplace contact and mask use are likely to contribute to differential COVID-19 risk. Across occupations, increasing workplace contact and reduced usage of face coverings presents an area of concern given ongoing high levels of community transmission and emergence of variants.

## Background

Severe Acute Respiratory Syndrome Coronavirus 2 (SARS-CoV-2) – the causative agent of the COVID-19 pandemic – spreads through populations via direct or indirect contact between individuals ^1,2^. Consequently, public health regulations aimed at reducing contact rates overall (e.g., ‘lockdowns’ and sectoral closures) and reducing effective contact where unavoidable (e.g., social distancing, face coverings) have been a cornerstone of the pandemic response worldwide.

Pandemic-related public health interventions have necessarily led to widespread and ongoing changes in the nature of work-related activities. Across several global regions, unprecedented numbers of workers who formerly attended in-person workplaces switched primarily or entirely to working from home or, where not possible to do so, relied on furlough schemes during extended periods of workplace closures ^3,4,5^. Workers in frontline roles have had to adapt to rapidly shifting mitigations, impacted by our evolving understanding of SARS-CoV-2 transmission, the underlying political and material context, and industry-related considerations ^5,6,7,8^. Balancing reopening workplaces with managing ongoing community transmission and the risk of SARS-CoV-2 variants presents an ongoing challenge. Occupation is consequently a fundamental and changing determinant of activity patterns for many people in the working age population, with consequent transmission-relevant implications for others with whom they interact within and outside of work.

Available contact surveys support the important role of work in determining contact rates across the pandemic. Despite large overall reductions in contacts across all settings during the initial months of the pandemic, surveys in the USA and UK suggested that work remained a persistent source of contacts in adults with a less dramatic decrease in contact rates than other locations ^9,10,11^. In the UK, adults who attended work during the pandemic had substantially higher mean contact rates than those who did not attend their workplace, with this pattern consistent but less pronounced across lockdown periods ^10^. While this initial evidence supports the key influence of work on contact patterns and consequently on potential SARS-CoV-2 exposure, investigation into indirect contact and mitigation in the workplace is also warranted to more thoroughly understand contact in this setting. Furthermore, investigation into workplace contact patterns across different occupations are lacking and are likely to vary substantially and influence risk.

The impact of pandemic-related interventions on different sectors, as well as pre-existing differences between occupations, likely influence the degree and routes of work-related SARS-CoV-2 exposure. Occupational differences in risk of infection, morbidity and mortality have emerged in both official statistics and research data from a variety of global regions ^12–20^, with patient or public-facing occupations and those requiring in-person attendance tending to demonstrate greater risk of infection and severe outcomes. The contribution of work-related exposure to differential risk is, however, difficult to measure and to delineate from non-work-related factors. Preliminary evidence from the UK suggests that contact at work partially mediates occupational differences in infection risk ^12,21^. However, understanding the mechanisms underlying differential infection risk by occupation is limited by a lack of data on how specific features of indirect and direct contact differ between occupations – i.e., in their frequency, duration, intensity, and mitigation in different workplace settings. As well as facilitating understanding of differential infection risk, investigation into differential features of occupational contact is relevant to inform both modelling of work-related transmission and to tailor public health interventions for specific occupational contexts.

The current study aimed to address this gap by quantifying workplace contact patterns by occupation across the second and third waves of the COVID-19 pandemic in England and Wales. Using electronic contact diaries completed as part of the Virus Watch cohort study ^22^ between November 2020 and November 2021, we set out to investigate how in-person workplace attendance and features of workplace contact, including intensity and duration of space sharing, direct contact, and wearing of face coverings, changed over time and differed between occupational groups.

## Methods

### Ethics Approval

The Virus Watch study was approved by the Hampstead NHS Health Research Authority Ethics Committee: 20/HRA/2320, and conformed to the ethical standards set out in the Declaration of Helsinki. All participants provided informed consent for all aspects of the study.

### Participants

Participants in the current study were an adult sub-cohort of the Virus Watch longitudinal cohort study enrolled prior to 09/11/2021 (*n*=50,759). Participants were included in the present study if they were:

1. an adult ≥16 years,
2. resident in England, for consistent timing of restrictions
3. were employed or self-employed full-time or part-time time and reported their occupation upon study registration,
4. and completed at least one contact diary survey between November 2020 and November 2021.

Further detail of the full Virus Watch cohort study, including inclusion criteria for the full cohort, can be obtained from the study protocol ^22^.

### Exposure

Participants’ entered their occupation as free text upon study registration, which we then assigned UK Standard Occupational Classification (SOC) 2020 ^23^ codes using semi-automatic processing in Cascot Version 5.6.3 ^24^. Occupations were subsequently classified into the following groups based on SOC codes (Supplementary Table S1; see ^12^ for further details of occupational classification): administrative and secretarial occupations; healthcare occupations; indoor trade, process & plant occupations; leisure and personal service occupations; managers, directors, and senior officials; outdoor trade occupations; sales and customer service occupations; social care and community protective services; teaching education and childcare occupations; transport and mobile machine operatives; and other professional and associate occupations (broadly office-based, non-essential professional occupations).

### Outcomes

All outcomes for this study were derived from electronic contact diaries delivered using REDCap ^25^, which prompted participants to select all settings where they spent time during a recent 24-hour period (between 5am on Monday and 5am Tuesday of the survey week). Workplace attendance was binary coded (yes/no) if they indicated attending their workplace outside of the home. Participants who attended work then responded to the following contact-related items, which were coded as follows in the current study to maximise group size while retaining theoretical significance:

- Maximum number of non-household members with whom the space was shared (regardless of distance): 0, 1-5, 6+ for work
- *If space was shared:* Total amount of time spent sharing space (work/transport) with others: <1 hour, 1-4 hours, 4+ hours for work
- *If space was shared:* Total number of close contacts at work (face-to-face contact within 1 metre, spending more than 15 minutes within 2 metres): 0, 1-5, 6+ close contacts
- *If any close contacts:* Frequency of wearing a face covering during close contact: binary for always wear (yes/no)

Diaries corresponded to the following dates, which reflected varying periods of legislation in England: 30 November 2020 (during second English national lockdown), 15 March 2021 (during third English national lockdown), 19 April 2021 (after restrictions on outdoor gatherings and non-essential retail relaxed), 24 May and 28 June 2021 (after restriction on indoor gatherings relaxed), 26 July 2021 (after most remaining COVID-19 restrictions relaxed), 29 September 2021 (after return to school and several months after relaxation of restrictions), and 23 November 2021 (further period after removal of restrictions and prior to concerns about transmission of the Omicron variant) ^6,7,8^

### Covariates

Where required (see Statistical Analyses), models were adjusted for the following covariates: age (<30, 30-39, 40-49, 50-59, 60+ years), sex at birth, employment status (full-time or part-time/other), shielding status (recommended to shield vs not), and vaccination status (unvaccinated, 1 dose, 2 doses, 3 doses). Only employment status and vaccination status were entered as time-varying covariates due to data availability.

### Statistical Analyses

We used logistic mixed models to investigate the effect of occupational group and time (diary month) on all contact-related outcomes (binomial logistic for workplace attendance and face covering usage; ordinal logistic for maximum number of people in shared workspace, time sharing workspace, and close contacts), including a random term to account for multiple submissions within individuals. To account for possible interactions between occupational group and time, we compared model fit before and after addition of an interaction term using likelihood ratio tests, with interactions included where *p*<0.05 for the likelihood ratio test; an interaction term was included in the final model for workplace attendance only.

In all models, the most prevalent occupational group - ‘Other professional and associate’ (see Results Table 1) – was set as the reference category. Following the UK Office for National Statistics ^21^, occupation-related results (main effects and interactions) were expressed as predictive probabilities rather than odds ratios to facilitate interpretation of differences between all occupational groups rather than comparison to the reference category.

**Table 1.**
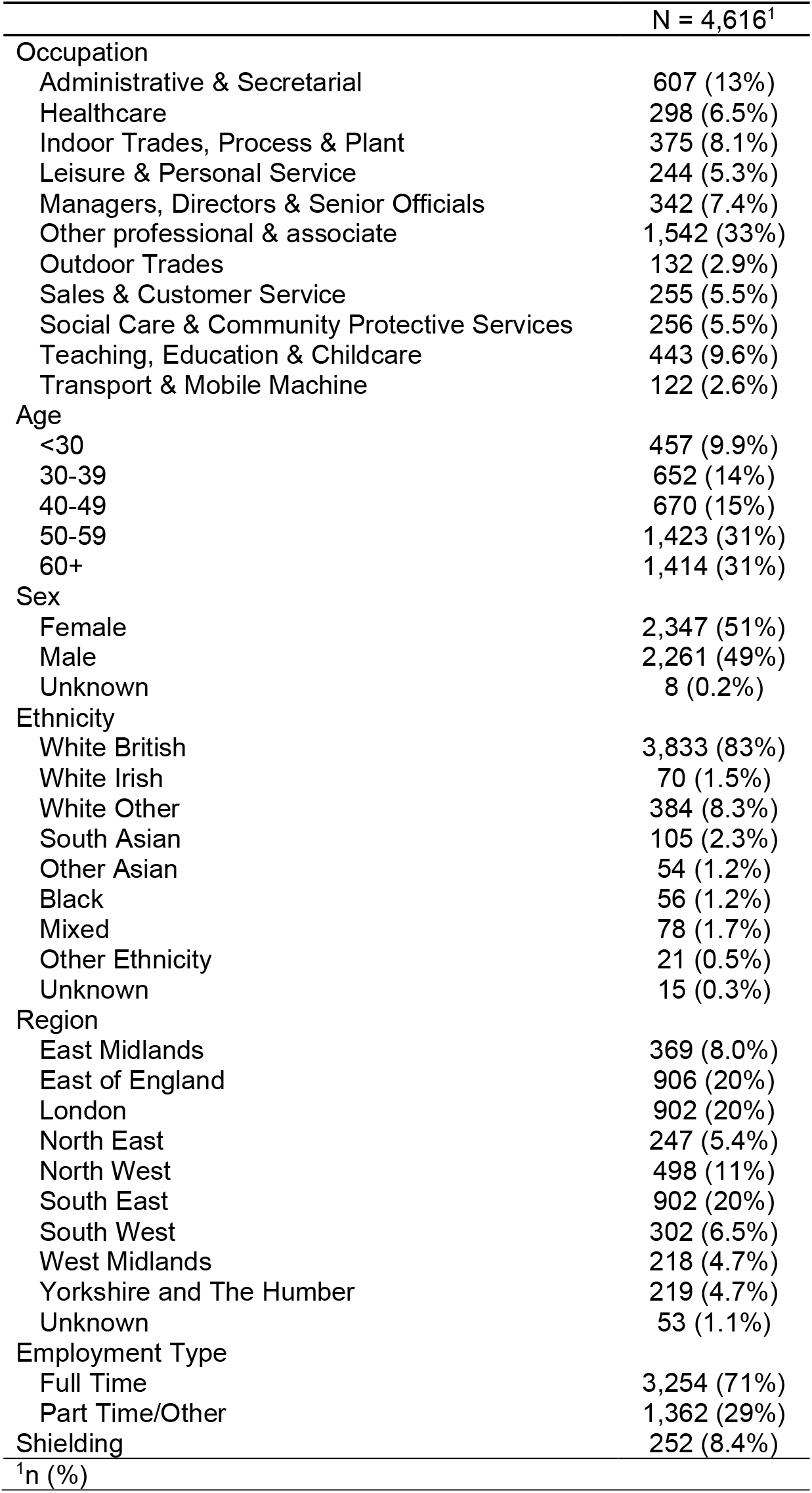
Demographic Features of Study Participants

|  | N = 4,616<sup>1</sup> |
| --- | --- |
| Occupation |  |
| Administrative & Secretarial | 607 (13%) |
| Healthcare | 298 (6.5%) |
| Indoor Trades, Process & Plant | 375 (8.1%) |
| Leisure & Personal Service | 244 (5.3%) |
| Managers, Directors & Senior Officials | 342 (7.4%) |
| Other professional & associate | 1,542 (33%) |
| Outdoor Trades | 132 (2.9%) |
| Sales & Customer Service | 255 (5.5%) |
| Social Care & Community Protective Services | 256 (5.5%) |
| Teaching, Education & Childcare | 443 (9.6%) |
| Transport & Mobile Machine | 122 (2.6%) |
| Age |  |
| <30 | 457 (9.9%) |
| 30-39 | 652 (14%) |
| 40-49 | 670 (15%) |
| 50-59 | 1,423 (31%) |
| 60+ | 1,414 (31%) |
| Sex |  |
| Female | 2,347 (51%) |
| Male | 2,261 (49%) |
| Unknown | 8 (0.2%) |
| Ethnicity |  |
| White British | 3,833 (83%) |
| White Irish | 70 (1.5%) |
| White Other | 384 (8.3%) |
| South Asian | 105 (2.3%) |
| Other Asian | 54 (1.2%) |
| Black | 56 (1.2%) |
| Mixed | 78 (1.7%) |
| Other Ethnicity | 21 (0.5%) |
| Unknown | 15 (0.3%) |
| Region |  |
| East Midlands | 369 (8.0%) |
| East of England | 906 (20%) |
| London | 902 (20%) |
| North East | 247 (5.4%) |
| North West | 498 (11%) |
| South East | 902 (20%) |
| South West | 302 (6.5%) |
| West Midlands | 218 (4.7%) |
| Yorkshire and The Humber | 219 (4.7%) |
| Unknown | 53 (1.1%) |
| Employment Type |  |
| Full Time | 3,254 (71%) |
| Part Time/Other | 1,362 (29%) |
| Shielding | 252 (8.4%) |
| <sup>1</sup>n (%) |  |

The November 2021 survey, comprising the most recent survey period and several months after the removal of most pandemic-related restrictions in England, was set as the reference category against which to compare various other time periods of restrictions. Time-related results expressed as odds ratios to facilitate comparison between different phases of restrictions.

Models were adjusted to account for plausible confounders of the relationship under investigation. Consequently, the model for workplace attendance was adjusted for age, sex, employment status, shielding, and vaccination status. Other sociodemographic factors were assumed to influence workplace attendance via their relationship with occupation and/or the factors controlled above, and consequently were not included in the model. Models for other features of workplace contact were limited to participants who attended work, and consequently these models were not adjusted as the effects of covariates on these factors were presumed to be mediated through occupation and workplace attendance. Missing data were sparse across included covariates (Table 1) and we performed complete case analysis.

## Results

A total of 4,616 participants submitted 23,762 contact diaries across the study period. Participants’ demographic features are reported in Table 1; participant selection is illustrated in Supplementary Figure S1. Vaccination status over time is illustrated in Supplementary Table S2.

Our mixed effect logistic regression models found that workplace attendance changed differentially over time between occupations, and that the number of people sharing the workspace, time spent sharing the workspace, number of workplace close contacts, and use of face coverings during close contact all varied significantly both by occupation and time period. (Figures 1-5)

**Figure 1.**
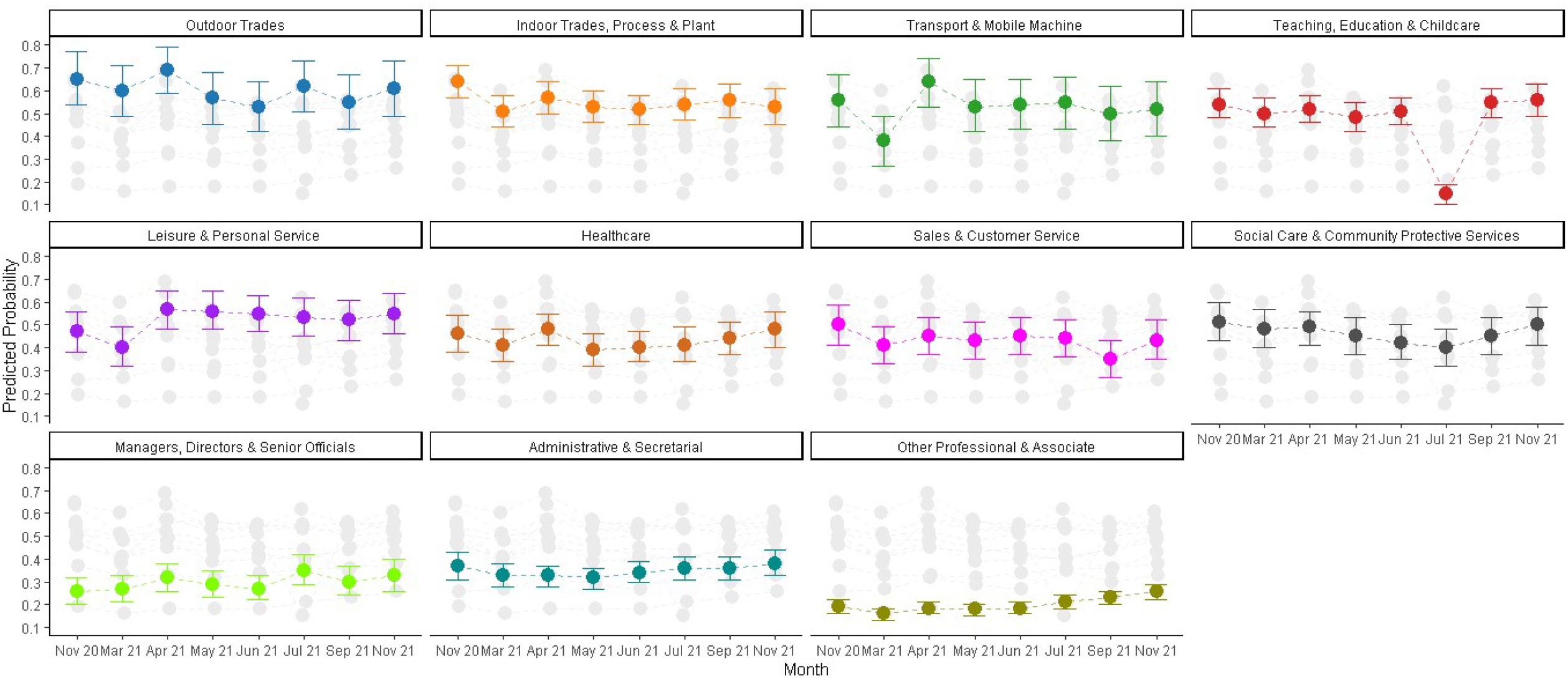
Interaction Plot (Occupational Group X Time) for Predicted Probability of Workplace Attendance on Survey Day

### Workplace Attendance (Figure 1)

Outdoor Trade occupations persistently exhibited the highest point estimates for predicted likelihood of workplace attendance on the diary day (0.55 [0.43,0.67] – 0.69 [0.59,0.79]), with confidence intervals overlapping with Indoor Trade occupations and Transport and Mobile Machine Operatives at all timepoints. For Leisure and Personal Service and Transport occupations, predicted probability of workplace attendance increased between March (respectively 0.40 [0.32, 0.49] and 0.38 [0.27, 0.49]) and April 2021 (respectively 0.57 [0.48, 0.65] and 0.64 [0.53, 0.74]) and remained relatively stable and high afterwards, possibly reflecting effects of sectoral re-openings. Predicted probabilities dropped considerably for Teaching, Education and Childcare occupations in July 2021 (0.15 [0.10, 0.19]) compared to previous and subsequent estimates (0.48 [0.42, 0.55] - 0.56 [0.49, 0.63]), in line with seasonal closures.

Other Professional and Associate occupations had the lowest predicted probability of workplace attendance at all timepoints (0.16 [0.13, 0.18] – 0.26 [0.22, 0.29]), with a trend towards increased in-person attendance from July 2021 onwards.

### Maximum Number of People in Workspace (Figure 2)

The predicted probability of sharing the workspace with more six or more others was highest for Teaching, Education and Childcare occupations (0.78 [0.75, 0.81]), Sales and Customer Service occupations (0.67 [0.62, 0.72]), and Healthcare occupations (0.64 [0.59,0.69]), exceeding estimates for all other occupational groups. Across all occupations, workspace sharing with either 1-5 or 6+ other people was more likely than no sharing at all. Outdoor trade occupations had the highest predicted probability of reporting no workspace sharing (0.30 [0.24,0.35]), exceeding all other groups.

**Figure 2.**
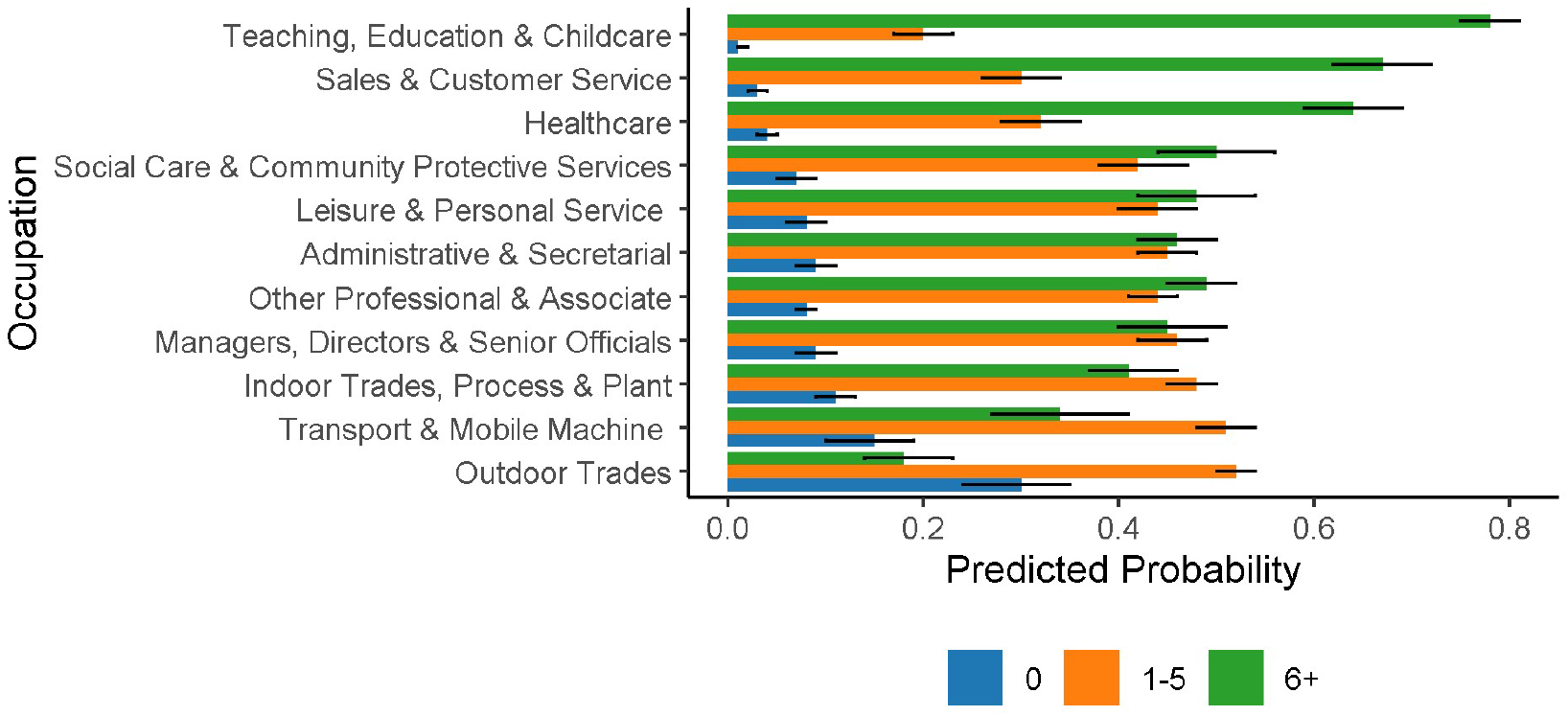
Predicted Probabilities for Maximum Number of People in Workspace by Occupational Group

Main effects of time for all workplace contact models are reported in Supplementary Table S3. Workers’ odds of workspace sharing across occupational groups were lower between November 2020 and July 2021 (OR range 0.47 [0.37-0.59] - 0.63 [0.49-0.79]) compared to November 2021. No evidence of a difference emerged between September and November 2021 (OR = 0.91 [0.72=1.15]).

### Time Spent Sharing Workspace (Figure 3)

Sharing the workspace for four or more hours was the most likely outcome across most occupational groups (predicted probability range 0.49 [0.43,0.55] – 0.62 [0.58,0.66]), Leisure and personal service, outdoor trade occupations and transport and mobile machine operatives - within which the most common occupations were large goods and delivery drivers (Supplementary Table S1) -had a relatively high likelihood of sharing the workspace with others for less than hour per day.

**Figure 3.**
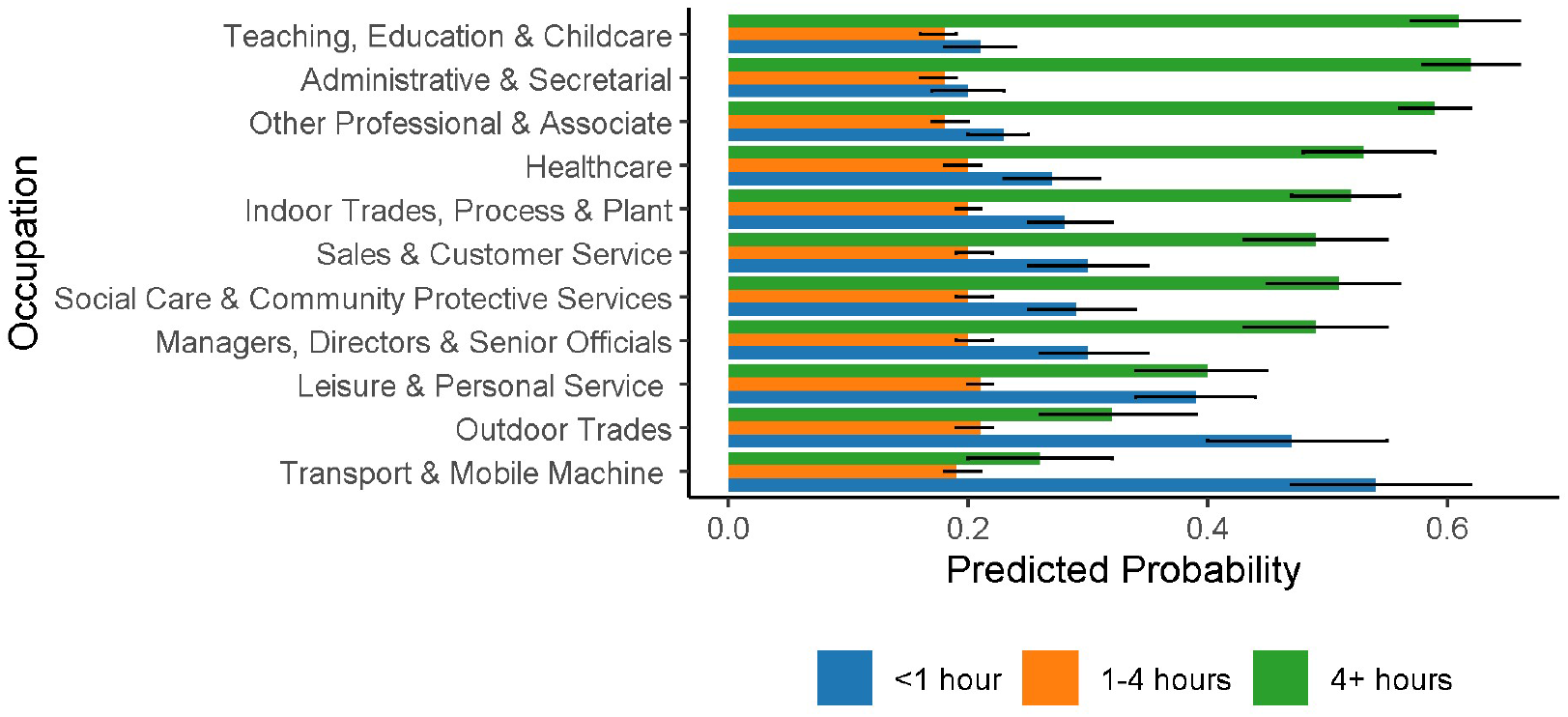
Predicted Probabilities for Time Spent Sharing Workspace by Occupational Group

The main effect of time indicated reduced length of time sharing the workspace in July (OR =0.68 [0.54,0.85]) only compared to November 2021 (Supplementary Table S3).

### Number of Close Contacts (Figure 4)

The predicted probability of having 6 or more contacts across the workday was highest for Teaching, Education and Childcare (0.36 [0.32,0.40]) occupations and Healthcare occupations (0.38 [0.33,0.43]). For all other occupational groups, the most likely number of close contacts across the workday was zero.

**Figure 4.**
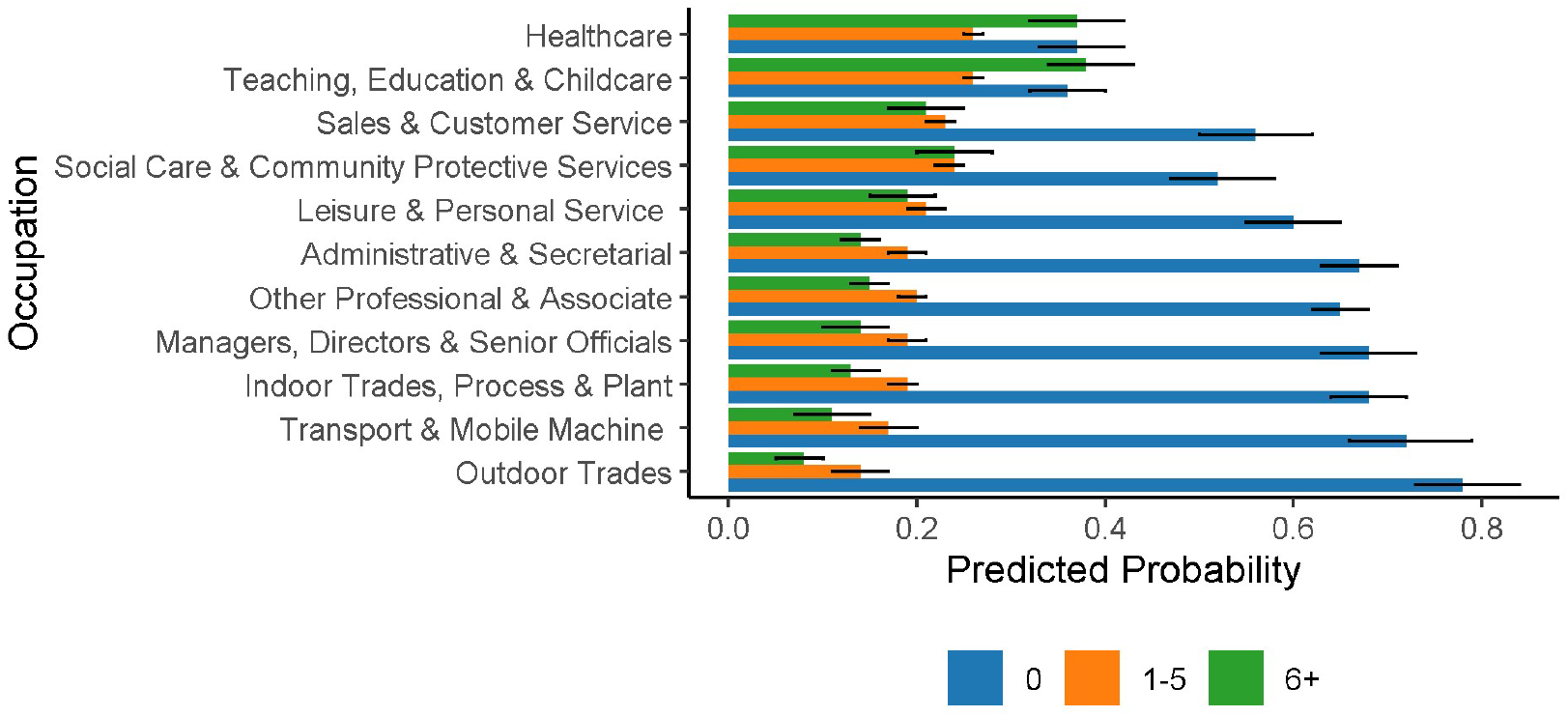
Predicted Probabilities for Number of Close Contacts at Work by Occupational Group

Across occupational groups, workers’ odds of reporting close contacts were lower between November 2020 and July 2021 (OR range 0.46 [0.37-0.57] - 0.65 [0.52-0.81]) compared to November 2021. No evidence of a significant difference emerged between September and November 2021 (OR = 0.89 [0.72=1.10]) (Supplementary Table S3).

### Wearing Face Covering during Close Contact (Figure 5)

Healthcare occupations had the highest predicted probability of wearing a face covering (0.90 [0.86,0.95]) with confidence intervals for the estimates overlapping only with Transport and Mobile Machine Operatives (0.79 [0.70,0.89]). There was considerable overlap between estimates for remaining occupations, with Outdoor Trades (0.34 [0.23, 0.45]), Other Professional and Associate occupations (0.51 [0.44, 0.57]), and Teaching, Education and Childcare Occupations (0.51 [0.44, 0.57]) demonstrating the lowest predicted probabilities.

**Figure 5.**
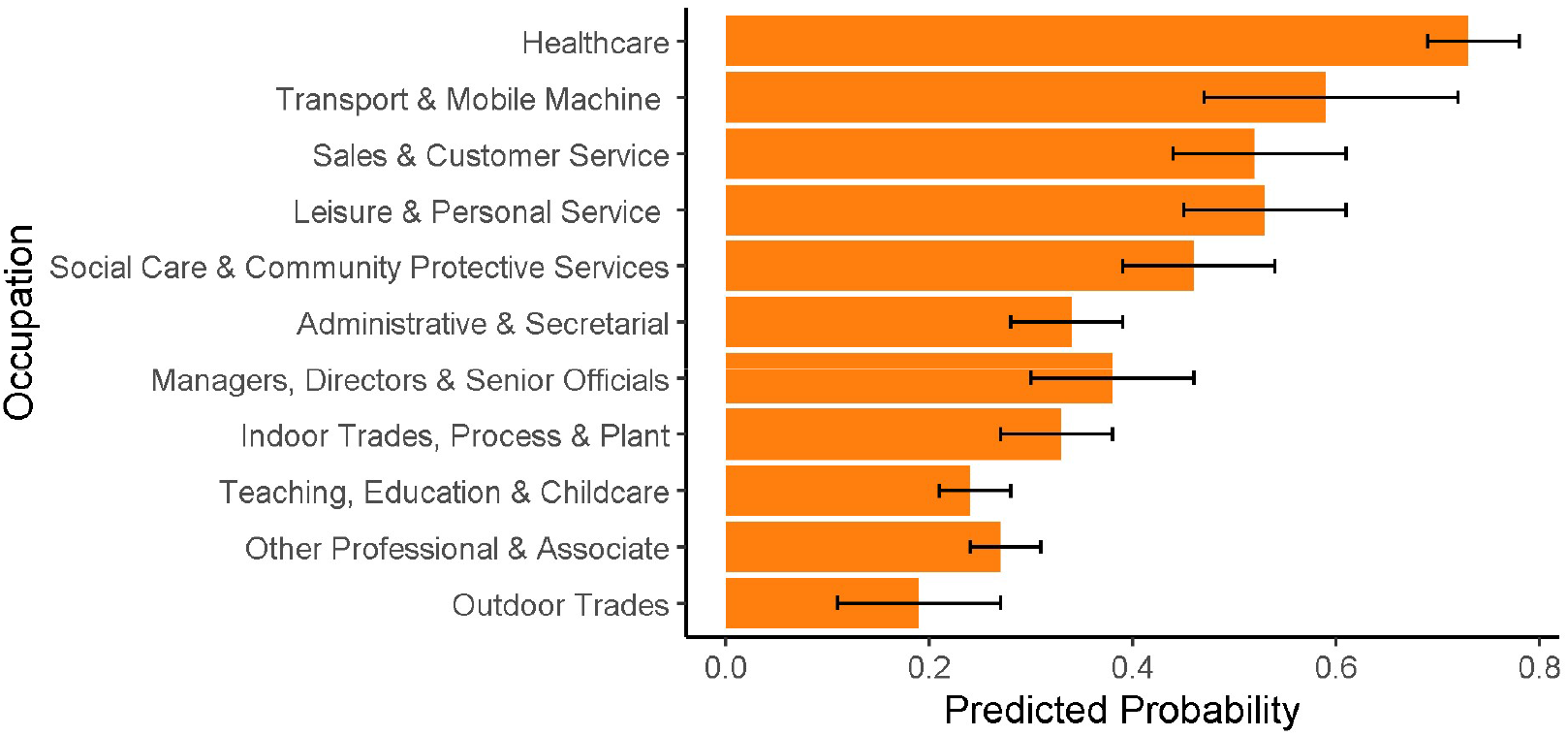
Predicted Probabilities for Wearing a Face Covering during Close Contact at Work by Occupational Group

All occupations had greater odds of wearing a face covering during close contact at work between November 2020 and July 2021 (OR range 2.64 [1.55, 4.48] - 20.71 [11.62, 36.90]) compared to November 2021. No substantial difference emerged between September and November 2021 (OR 0.87 [0.51,1.47]) (Supplementary Table S3).

## Discussion

### Key Findings and Interpretation

This study aimed to investigate in-person workplace attendance and workplace contact patterns by occupation across the second and third waves of the COVID-19 pandemic in England. Differential patterns of in-person workplace attendance and features of workplace contact were identified across occupational groups, with variation in at-risk groups depending on the characteristic under investigation. Across all occupations, intensity of workspace sharing and close contact increased after September 2021 and likelihood of wearing a face covering during close contact decreased. This shift corresponds to the periods after which most public health measures, including mandates around social distancing and mask wearing in some public spaces, had been lifted in England despite high ongoing levels of community transmission ^6,7^.

Probability of workplace attendance changed differentially over time between occupational groups, broadly in line with previous classifications for frontline versus non-frontline roles ^26^. Trade and transport occupations demonstrated high probability of in-person attendance, as did leisure/service and teaching occupations in line with periods of sectoral opening. Conversely, Other Professional and Associate occupations - broadly comprising non-essential, office-based roles - persistently demonstrated the lowest likelihood of in-person workplace attendance with a high proportion continuing to work mainly at home even after lifting of restrictions.

Occupational sectors identified in previous studies and surveillance data as having elevated risk of SARS-CoV-2 infection - largely essential and public-facing occupations including healthcare, transportation, education, indoor trade and service occupations ^12–20^ - also tended to demonstrate high likelihood of workplace attendance and/or multiple elements of workplace contact-related risk in the current study. Notably, workers in teaching/education/childcare occupations demonstrated high probabilities of workplace attendance, workspace sharing and close contact, as well as relatively low probability of wearing a face covering during close contact. While workspace sharing and close contact may be difficult to avoid in many educational roles, uptake of face coverings as well as potential environmental mitigations addressing indirect and direct contact-related risk may be particularly beneficial in these environments. Healthcare workers also tend to demonstrate elevated infection risk based on research and surveillance data, particularly during the first pandemic wave ^20,27–30^. In the present study, they also demonstrated more intense space sharing and close contact than most other groups. While it could not be directly assessed in the present study, access to personal protective equipment and other mitigation methods including prioritisation for vaccination was likely instrumental in mitigating contact-related risk in later pandemic waves.

Despite observed variation in features of workplace contact across occupational groups, workspaces tended to be shared with others for long periods of time across most occupations. Probability of close contact across occupations was lower than that of space sharing, but reporting may be particularly influenced by social desirability bias - close contact was a target of public health messaging in England during the pandemic - as well as the stringent definition of contact. While overall probabilities of close contact may have consequently been underestimated, this is unlikely to have influenced between-occupational differences. Furthermore, indirect contact can still present a transmission risk, particularly in high-footfall, poorly ventilated indoor environments ^1,2,31^. In light of ongoing high levels of SARS-CoV-2 transmission and emergence of new variants, increasing workspace sharing and close contact across occupations with reduced usage of face coverings is likely a major contributor to transmission. Public health interventions to reduce the number of individuals sharing workspaces - including promoting working from home where possible - and to promote the uptake of mitigation methods such as face coverings are important measures to slow transmission.

### Strengths and Limitations

Strengths of this study include the large, diverse cohort that allowed us to investigate workplace contact across a range of occupational groups. Repeated surveys covered key periods of the second and third pandemic waves in England, and were repeated after major changes in pandemic-related restrictions over time.

Several important limitations, however, should be considered in interpreting these findings. The study cohort is not representative of the English population. Both occupation and contact patterns were measured in broad categories. Occupational groups are likely to include specific roles with different risk profiles, but we lacked power to investigate in further detail. Notably, contact patterns amongst the Transport and Mobile Machine operative group may have been influenced by the relatively large proportion of large goods and delivery drivers relative to public transport workers; however, we were unable to disaggregate these occupations further. Self-reported contact and activities may have been impacted by recall bias and social desirability bias, particularly during periods of stringent restrictions. Findings are not generalisable to the first pandemic wave when many infections may have occurred, particularly in some frontline occupational groups such as health and social care workers ^20,27–29^. Linking these findings directly to infection risk was beyond the scope of the present study.

Each contact survey related to a single weekday in order to facilitate detailed recall of activities, but may not have been representative of a normative weekday for participants. For participants with hybrid or part-time working patterns, the selection of Monday may have influenced the likelihood of in-person workplace attendance as mid-week workplace working may be more common. Due to the survey design, it was not possible to attribute a specific reason for workplace non-attendance, which may have been driven by workplace closure, self-isolation, or other reasons for non-attendance including those independent of the pandemic. Estimates of in-person workplace attendance may have been systematically biased downwards during some timepoints, particularly July 2021 when high levels of community transmission led to a surge in requests to self-isolate via the English National Health Service Test and Trace mobile application ^32^; these requests may have affected some public-facing occupations differentially. Further details around environmental features of the workspace and mitigation methods used at work were beyond the scope of this survey. In particular, the use of personal protective equipment (PPE) and ventilation in indoor spaces could not be assessed, and are relevant moderators of contact-related risk. It was also not possible to attribute the source of close contact or space sharing (i.e., whether this was driven by colleagues and/or the public).

## Conclusions

Our findings provide quantitative evidence of differential workplace attendance and workplace contact patterns by occupation during the COVID-19 pandemic. This study also demonstrates change over time in workplace contact across the pandemic, with evidence of greater degree of space sharing and close contact at work and lower probability of wearing a face covering during close contact after the lifting of relevant national public health measures. These findings provide preliminary evidence around variation in risk-relevant features of workplace contact to inform further research and risk mitigation of SARS-CoV-2 and other respiratory infections in the workplace.

## Supporting information

STROBE Checklist

## Data Availability

We aim to share aggregate data from this project on our website and via a "Findings so far" section on our website - https://ucl-virus-watch.net/. We will also be sharing individual record level data on a research data sharing service such as the Office of National Statistics Secure Research Service. In sharing the data we will work within the principles set out in the UKRI Guidance on best practice in the management of research data. Access to use of the data whilst research is being conducted will be managed by the Chief Investigators (ACH and RWA) in accordance with the principles set out in the UKRI guidance on best practice in the management of research data. We will put analysis code on publicly available repositories to enable their reuse.

