## Supplementary Materials for "Workplace Contact Patterns in England during the COVID-19 Pandemic: Analysis of the Virus Watch prospective cohort study"

### Occupational Classification

**Supplementary Table S1. UK Standard Occupational Classification 2020 (SOC-2020) Codes within Virus Watch Occupational Categories**

| Virus Watch Occupational Category | UK SOC-2020 Codes | Three Most Prevalent Occupations in Study Sample* (SOC-2020 Unit Group) |
|---|---|---|
| Administrative & Secretarial Occupations | 4111-4217, 9211, 9219, 9233 | 1. Other administrative occupations n.e.c. (20%, n= 122)<br>2. Book-keepers, payroll managers, and wage clerks (9.4%, n=57)<br>3. Personal assistants and other secretaries (7.7%, n=47) |
| Healthcare Occupations | 2211-2259, 3211-3219, 3240, 6131-6133 | 1. Other nursing professionals (19%, n=56)<br>2. Generalist medical practitioners (8.4%, n=25)<br>3. Nursing auxiliaries and assistants (8.1%, n=24) |
| Indoor Trades, Process & Plant Occupations | 5211-5250, 5315-5317, 5321-5323, 5411-5449, 8111-8149, 8160, 9131-9139, 9241-9259 | 1. Warehouse operatives (10%, n=38)<br>2. Metalworking production and maintenance fitters (8.8%, n=33)<br>3. Electricians and electrical fitters (7.5%, n=28) |
| Leisure & Personal Service Occupations | 1221-1225, 1252, 1253, 1256, 1257, 6121, 6129, 6211-6250, 9221-9229, 9231, 9261-9269 | 1. Cleaners and domestics (15%, n=37)<br>2. Kitchen and catering assistants (9.8%, n=24)<br>3. Hairdressers and barbers (8.6%, n=21) |

| | | |
|---|---|---|
| Managers, Directors & Senior Officials | 1111-1161, 1171,1172, 1211, 1212, 1231, 1241-1243, 1251, 1254, 1255, 1258, 1259 | 1. Financial managers and directors (15%, n=50)<br>2. Functional managers and directors n.e.c. (10%, n=35)<br>3. Marketing, sales, and advertising directors (10%, n=35) |
| Other Professionals & Associate Professionals | 2111-2162, 2411-2455, 2471-2494, 3111-3133, 3411-3582 | 1. Programmers and software development professionals (6.5%, n=100)<br>2. Business and financial project management professionals (4.2%, n=64)<br>3. Management consultants and business analysts (3.8%, n=59) |
| Outdoor Trade Occupations | 5111-5119, 5311-5314, 5319, 5330, 8151-8159, 9111- 9129 | 1. Gardeners and landscape gardeners (21%, n=28)<br>2. Construction and building trades n.e.c. (17%, n=23)<br>3. Construction operatives n.e.c. (12%, n=16) |
| Sales & Customer Service Occupations | 7111-7220 | 1. Sales and retail assistants (35%, n=88)<br>2. Customer service occupations n.e.c. (11%, n=29)<br>3. Retail cashiers and check-out operators (11%, n=29) |
| Social Care & Community Protective Services | 1162, 1163, 1232, 2461-2469, 3221-3229, 3311-3319, 6134-6138, 6311-6312 | 1. Care workers and home carers (24%, n=61)<br>2. Welfare and housing associate professionals n.e.c. (12%, n=30)<br>3. Youth and community workers (9%, n=23) |
| Teaching, Education & Childcare Occupations | 1233, 2311-2329, 3231, 3232, 6111-6117, 9232 | 1. Higher education teaching professionals |

| | | |
|---|---|---|
| | | (17%, n=75)<br>2. Education advisers and school inspectors (16%, n=70)<br>3. Secondary education teaching professionals (13%, n=56) |
| Transport & Mobile Machine Operatives | 8211-8239 | 1. Large goods vehicle drivers (22%, n=27)<br>2. Delivery drivers and couriers (19%, n=23)<br>3. Bus and coach drivers (15%, n=18) |

**Abbreviations:** n.e.c. = not elsewhere classified; * Limited to three most prevalent occupations per category to prevent declarative disclosure and due to large number of occupations across sample (*n*=381)

**Supplementary Figure S1. Flow Diagram of Participant Inclusion**

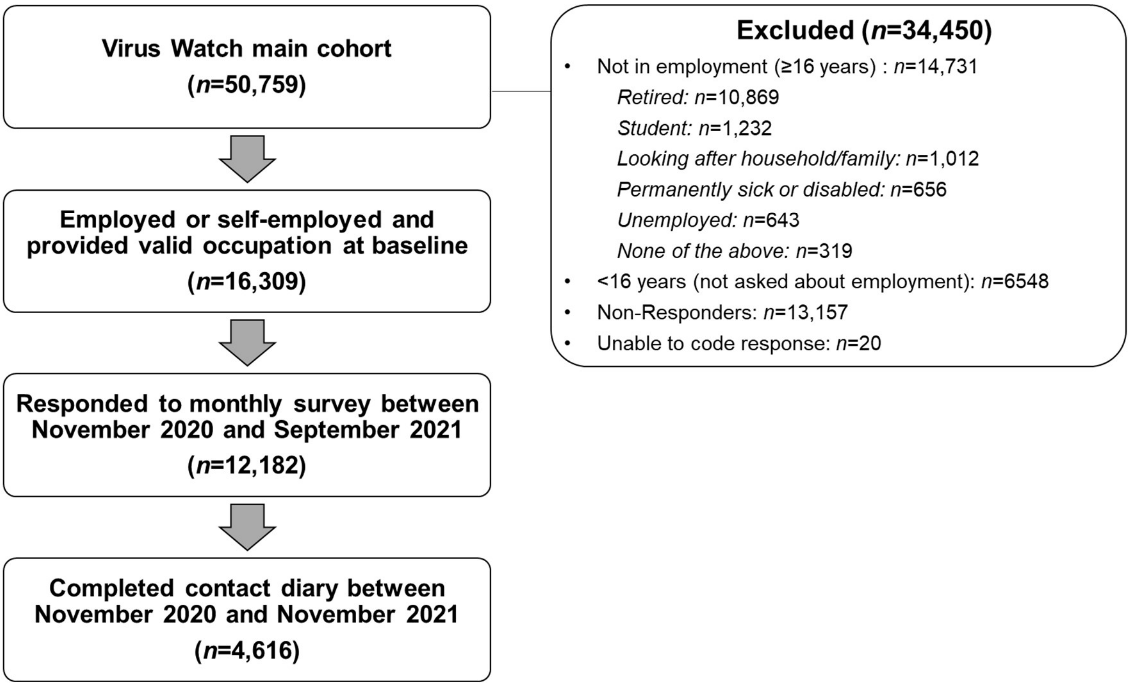

**Supplementary Table S2. Participant Vaccination Status over Time**

| | **Total –** n (row %) | **Unvaccinated** - n (row %) | **One Dose** – n (row %) | **Two Doses** - n (row %) | **Three Doses** – n (row %) |
|---|---|---|---|---|---|
| **November 2020** | 3,049 (100%) | 3,049 (100%) | 0 (0%) | 0 (0%) | 0 (0%) |
| **March 2021** | 3,397 (100%) | 1,440 (42%) | 1,859 (55%) | 98 (2.9%) | 0 (0%) |
| **April 2021** | 3,361 (100%) | 795 (24%) | 1,938 (58%) | 628 (19%) | 0 (0%) |
| **May 2021** | 3,144 (100%) | 393 (12%) | 1,013 (32%) | 1,738 (55%) | 0 (0%) |
| **June 2021** | 2,989 (100%) | 138 (4.6%) | 536 (18%) | 2,315 (77%) | 0 (0%) |
| **July 2021** | 2,863 (100%) | 102 (3.6%) | 335 (12%) | 2,426 (85%) | 0 (0%) |
| **September 2021** | 2,538 (100%) | 63 (2.5%) | 142 (5.6%) | 2,259 (89%) | 74 (2.9%) |
| **November 2021** | 2,421 (100%) | 42 (1.7%) | 80 (3.3%) | 1,151 (48%) | 1,148 (47%) |

**Supplementary Table S3. Main Effects of Time for Workplace Contact Models**

| | **Number of People in Workspace –** OR (95% CI) | **Time Spent Sharing Workspace –** OR (95% CI) | **Number of Workplace Close Contacts–** OR (95% CI) | **Face Covering During Close Contact –** OR (95% CI) |
|---|---|---|---|---|
| **November 2020** | 0.53 (0.42, 0.67) | 1.12 (0.90, 1.40) | 0.64 (0.51, 0.79) | 4.56 (2.67, 7.78) |
| **March 2021** | 0.47 (0.37, 0.59) | 0.88 (0.71, 1.10) | 0.46 (0.37, 0.57) | 20.71 (11.62, 36.90) |
| **April 2021** | 0.58 (0.46, 0.73) | 0.93 (0.76, 1.15) | 0.50 (0.41, 0.61) | 13.16 (7.62, 22.73) |
| **May 2021** | 0.61 (0.48, 0.76) | 0.86 (0.70, 1.07) | 0.56 (0.45, 0.69) | 5.90 (3.46, 10.06) |
| **June 2021** | 0.63 (0.49, 0.79) | 0.90 (0.72, 1.12) | 0.53 (0.43, 0.66) | 2.64 (1.55, 4.48) |
| **July 2021** | 0.56 (0.44, 0.71) | 0.68 (0.54, 0.85) | 0.65 (0.52, 0.81) | 2.69 (1.57, 4.61) |
| **September 2021** | 0.91 (0.72, 1.15) | 0.88 (0.70, 1.09) | 0.89 (0.72, 1.10) | 0.87 (0.51, 1.47) |
| **November 2021** | ref | ref | ref | ref |

Note: OR = odds ratio; CI = confidence intervals
